# Vaginal bacterial load in the second trimester is associated with early preterm birth recurrence: a nested case-control study

**DOI:** 10.1101/2021.01.14.21249680

**Authors:** Laura Goodfellow, Marijn C. Verwijs, Angharad Care, Andrew Sharp, Jelena Ivandic, Borna Poljak, Devender Roberts, Christina Bronowski, A. Christina Gill, Alistair C. Darby, Ana Alfirevic, Bertram Muller-Myhsok, Zarko Alfirevic, Janneke H.H.M. van de Wijgert

## Abstract

**Objective:** To assess the association between vaginal microbiome (VMB) composition and recurrent early spontaneous preterm birth (sPTB)/preterm prelabour rupture of membranes (PPROM).

**Design:** Nested case-control study.

**Setting:** UK tertiary referral hospital.

**Sample:** High-risk women with previous sPTB/PPROM <34^+0^ weeks gestation who had a recurrence (n=22) or delivered at ≥37^+0^ weeks without PPROM (n=87).

**Methods:** Vaginal swabs collected between 15-22 weeks gestation were analysed by 16S rRNA gene sequencing and 16S quantitative PCR.

**Main outcome measure:** Recurrent early sPTB/PPROM.

**Results:** 28/109 high-risk women had anaerobic vaginal dysbiosis, with the remainder dominated by lactobacilli (*L. iners* 36/109, *L. crispatus* 23/109, or other 22/109). VMB type, diversity, and stability were not associated with recurrence. Women with a recurrence, compared to those without, had a higher median vaginal bacterial load (8.64 vs. 7.89 log_10_ cells/μl, adjusted odds ratio (aOR)=1.90, 95% confidence interval (CI)=1.01-3.56, p=0.047) and estimated *Lactobacillus* concentration (8.59 vs. 7.48 log_10_ cells/μl, aOR=2.35, CI=1.20-4.61, p=0.013). A higher recurrence risk was associated with higher median bacterial loads for each VMB type after stratification, although statistical significance was reached only for *L. iners*-domination (aOR=3.44, CI=1.06-11.15, p=0.040). Women with anaerobic dysbiosis or *L. iners*-domination had a higher median vaginal bacterial load than women with a VMB dominated by *L. crispatus* or other lactobacilli (8.54, 7.96, 7.63, and 7.53 log_10_ cells/μl, respectively).

**Conclusions:** Vaginal bacterial load is associated with early sPTB/PPROM recurrence. Domination by lactobacilli other than *L. iners* may protect women from developing high bacterial loads. Future PTB studies should quantify vaginal bacteria and yeasts.

**Funding:** Wellbeing of Women, London, UK

**Tweetable abstract:** Increased vaginal bacterial load in the second trimester may be associated with recurrent early spontaneous preterm birth.

## Introduction

Previous spontaneous preterm birth (sPTB) or preterm prelabour rupture of membranes (PPROM) is the strongest risk factor for recurrent sPTB/PPROM.^1^ This may implicate a genetic contribution^2^ but does not rule out a contribution of persistent microbes and/or inflammation in the female reproductive tract.^3^

16S ribosomal RNA (16S rRNA) sequencing was first used to assess the relationship between the bacterial vaginal microbiota (VMB) and PTB in 2014.^4^ Since then over 2000 pregnant women have been assessed in 12 studies (Appendix A, Table A1). The majority of these studies suggested a contribution of the VMB composition to PTB, but results have been inconsistent. The healthy vagina in pregnancy is dominated by *Lactobacillus* species but contains up to 80 different *Lactobacillus* and other bacterial taxa, and the optimal ways of summarising and categorising sequencing data to determine clinically relevant associations with PTB are yet to be determined.^5,6^ Studies to date have shown that increased relative abundances of *L. crispatus, L. gasseri*, and *L. jensenii* were either not associated with PTB or associated with a reduced PTB risk, and increased relative abundances of bacterial vaginosis (BV)-associated anaerobes were either not associated with PTB or associated with an increased PTB risk (Table A1). Results for *L. iners* relative abundance were inconsistent, with 6 studies showing no association, two studies an increased risk, and one study a decreased risk (Table A1). Importantly, all studies except two^7,8^ relied on compositional data, meaning that an increase in the proportion of one taxon always leads to a decrease in the proportions of other taxa.^9^ Furthermore, an increase in a proportion does not necessarily indicate an increase in absolute quantity. VMB researchers have only recently begun to semi-quantify 16S rRNA sequencing data by combining sequencing data with a quantitative PCR (qPCR) of the 16S rRNA gene.^10^ The two studies to date that included some quantification reported opposite results, with one reporting a positive association between overall vaginal bacterial load and PTB risk,^7^ and the other a negative association.^8^

Our aim was to assess associations between VMB compositions and recurrent early sPTB/PPROM (delivery at <34^+0^ weeks compared to delivery at ≥37^+0^ weeks) among high-risk pregnant women who had had a previous early sPTB/PPROM, using compositional and semi-quantitative 16S sequencing data.

## Methods

### Study design and population

Two cohorts of women at high and low-risk of PTB were enrolled at Liverpool Women’s Hospital, UK, from 1 June 2015 until 31 December 2017. The hospital’s patient and public engagement group helped guide the research and recruitment (Appendix A). High-risk was defined as a history of sPTB or PPROM at 16^+0^-33^+6^ weeks gestation, and low-risk as having had at least one previous term birth (≥37^+0^ weeks) without PPROM and never having had a PTB or PPROM. In the low-risk cohort, but not in the high-risk cohort, women were excluded if they had had significant cervical surgery or previous obstetric or medical problems (Appendix A). For this nested case-control study, we selected 109 women from the high-risk cohort (N=137) and 145 women from the low-risk cohort (N=223) based on the pregnancy outcome (sPTB or PPROM 16^+0^-33^+6^ weeks or birth ≥37^+0^ weeks in the high-risk cohort and birth ≥39^+0^ weeks in the low-risk cohort) and availability of valid VMB data (Figure 1). Birth outcomes were classified independently by two obstetricians experienced in PTB management. When there was a discrepancy between them, the case was reviewed by a third experienced obstetrician until the team reached consensus. Participants with medically indicated PTBs, or for whom it was unclear whether a birth was truly spontaneous, were excluded. We used relevant parts of the CROWN initiative core outcome set for PTB research^11^ (Appendix A).

**Figure 1:**
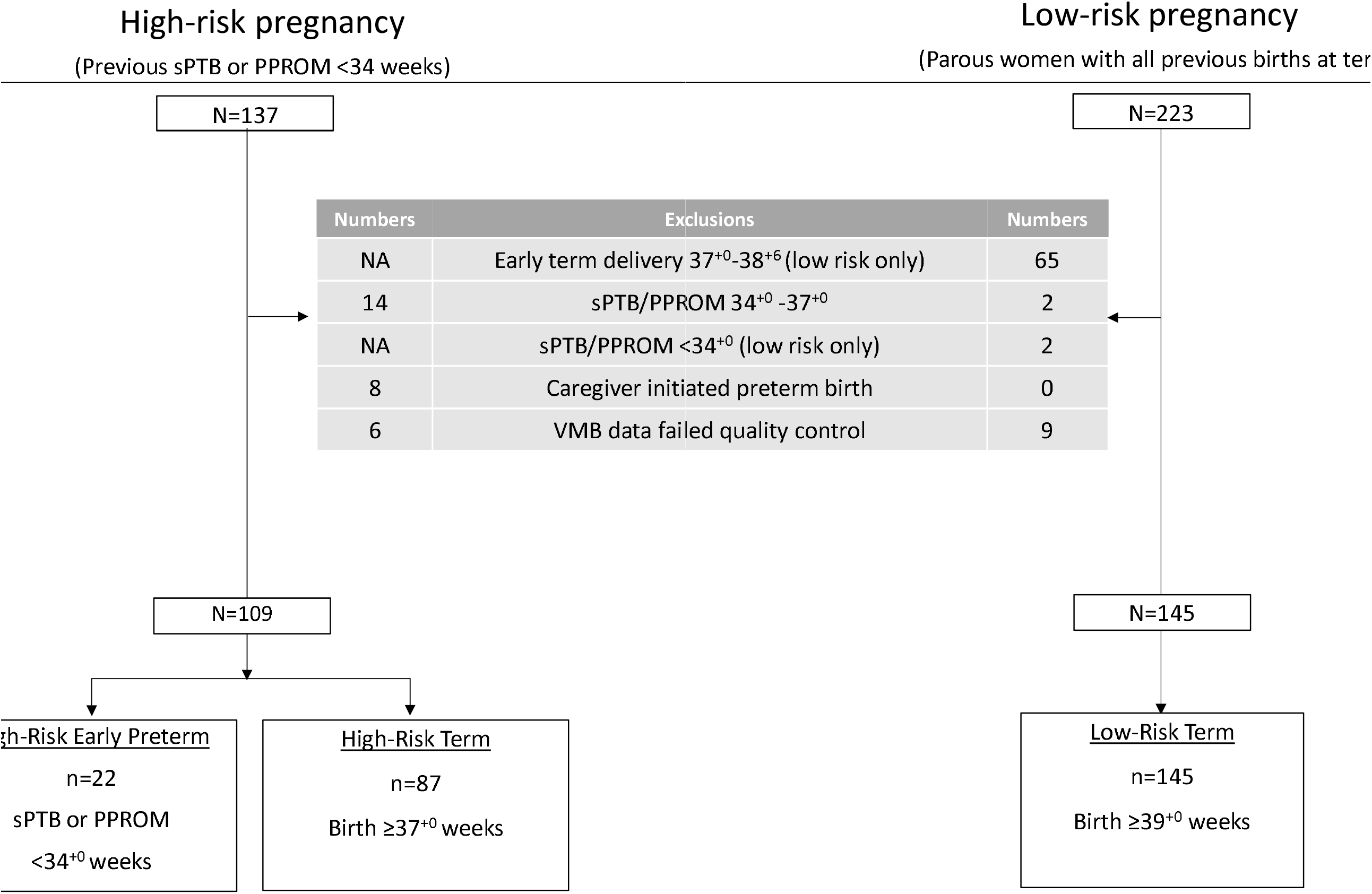
Participant flow.

### Clinical procedures

Participants were invited to two study visits at approximately 16 weeks (15^+1^-18^+6^ weeks) and 20 weeks (19^+0^-23^+0^) gestation. At each visit, a study obstetrician interviewed the participant; collected high vaginal swabs during a speculum examination for VMB assessments at the University of Liverpool Center for Genomic Research (HydraFlock standard tapered swabs, Medical Wire and Equipment, Corsham, UK) and for quantitative assessment of foetal fibronectin and culture by the local NHS laboratory; determined cervical length; and did a 3D foetal ultrasound. All other procedures during pregnancy, and all treatments, were in accordance with UK national guidelines.^12^ Preterm birth prevention therapy (cerclage, vaginal pessary, or vaginal progesterone) was offered if cervical length was ≤ 25mm or based on clinician and patient preference in case of a large change in cervical length between measurements. None of the low-risk women required an intervention but 71.6% (78/109) of the high-risk women did (Table 1). Initiation of this treatment was always after the VMB sampling visits.

**Table 1:**
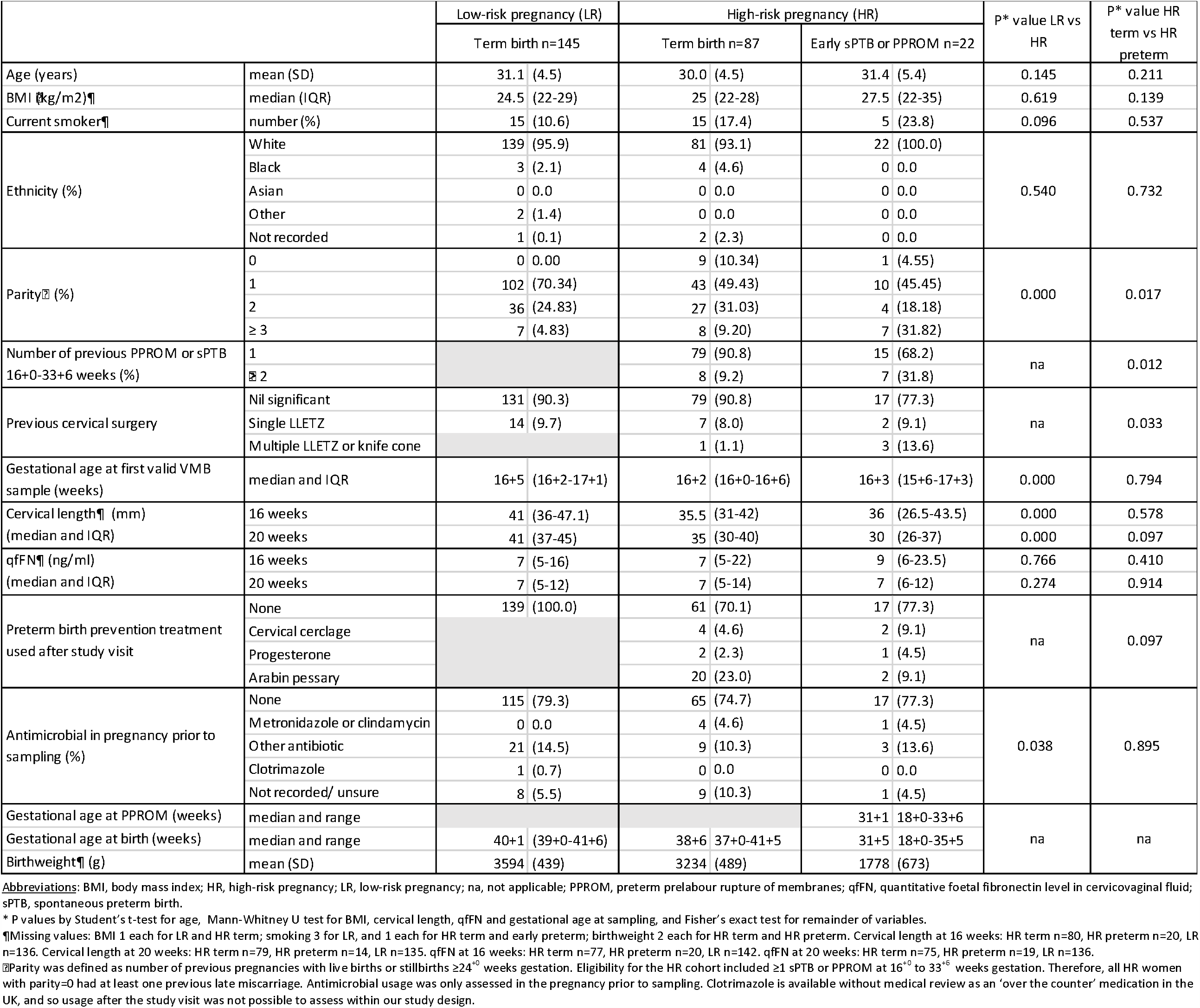
Participant characteristics by pregnancy risk group and outcome.

### VMB laboratory assessments

DNA was extracted from one swab per woman per visit using lysozyme lysis and bead-beating procedures combined with the Qiagen DNeasy Blood and Tissue kit (Qiagen, Manchester, UK) (Appendix A).^13^ The V3-V4 region of the 16S rRNA gene was amplified and sequenced on an Illumina HiSeq 2500 instrument (Illumina, San Diego, CA) run in rapid mode, 2×300bp using a 250PE and 50E kit. The panbacterial 16S rRNA gene copy concentration per sample was determined using the BactQuant quantitative PCR (qPCR) assay.^14^ Molecular data processing steps are described in Appendix A.

We only used the first set of valid sequencing results for each participant in the analyses because of the short time period between the two study visits, with one exception. In the subset of participants with valid sequencing results for both visits (88 high-risk and 129 low-risk women) we used data from both visits for the VMB stability analyses.

### Creation and selection of VMB variables

#### Existing approaches

We systematically reviewed the published literature to identify VMB variables that were associated with PTB in at least one study (Appendix A, Table A1). These included: richness and Simpson diversity (1-D) as continuous variables; two categorical VMB composition variables with each participant assigned to one category (the community state types (CSTs) described by Ravel et al^15^ and *Lactobacillus* groups (dominant, intermediate, and deplete) based on *Lactobacillus* relative abundance);^16^ stability groups, slightly modified from Romero *et al*;^17^ and the presence/absence of specific taxa of interest (listed in Table A1).

### New approaches

The final three sets of VMB variables are based on our previous work and were applied to PTB research for the first time.^18–21^ The first is a categorical VMB composition variable that we named ‘VMB type’. These VMB types are similar to Ravel’s CSTs^15^ but with an increased emphasis on the relative abundance of pathobionts (Appendix A). We defined pathobionts as bacteria that are considered more pathogenic than BV-anaerobes and that often co-occur with lactobacilli instead of BV-anaerobes, such as streptococci. Secondly, each non-minority amplicon sequence variant (ASV; equivalent to a taxon) in each sample was allocated to one of four ‘bacterial groups’ based on the published literature (Appendix B): lactobacilli; BV-anaerobes; pathobionts; and a rest group of ‘other bacteria’.^20^ Within each sample, read counts of ASVs belonging to the same bacterial group were summed. This resulted in four continuous relative abundance variables (one for each bacterial group) per sample, which sum to 1.0 for each sample. Finally, we converted these bacterial group relative abundances into estimated concentrations (again four continuous variables, one for each bacterial group) making use of the BactQuant results (Appendix A). The BactQuant data were also used to determine estimated concentrations of taxa of interest (listed in Table A1) and the total bacterial load in each sample.

### Statistical analysis

Our primary comparisons were between high-risk women who gave birth ≥37^+0^ weeks gestation and high-risk women who had a recurrent early sPTB or PPROM <34^+0^ weeks. Low-risk women who delivered at ≥39^+0^ weeks gestation without PPROM were used as the normal reference group because we had never before characterised VMB compositions in pregnant women residing in Liverpool. Characteristics between these three groups were compared by student’s t-test for age, Mann-Whitney U test for other continuous variables, and Fisher’s exact test for binary and categorical variables. The various VMB variables described above were compared between the two high-risk groups using unadjusted and adjusted logistic regression, with adjustments for body mass index (BMI; as a quadratic term due to the bimodal association with PTB), history of cervical surgery (none, single LLETZ, or multiple LLETZ or knife cone biopsy), and smoking at the time of enrolment (yes/no). Quartiles of the expected distributions of total vaginal bacterial load and total estimated *Lactobacillus* concentration were generated using data from the low-risk reference group, and high-risk participants were allocated to one of these quartiles. These newly created categorical variables were also compared between the two high-risk groups of interest by logistic regression. Finally, in an effort to differentiate between the effects of total vaginal bacterial load and the types of bacteria that make up this load, women were stratified by VMB type, and the logistic regression analyses were repeated for each stratum.

## Results

We recruited 137 high-risk women and 223 low-risk women. Of 131 high-risk women with VMB data that passed quality control, 22 had a recurrent early sPTB/PPROM and 87 had term births ≥37^+0^ weeks without PPROM (Figure 1); the remaining women were excluded because they delivered between 34^+0^ and 37^+0^ weeks or had a birth initiated by a caregiver. Of 214 low-risk participants with VMB data that passed quality control, 145 gave birth at ≥39^+0^ weeks without PPROM and were used as the normal reference group.

The participant characteristics of the three groups were similar (Tables 1 and A2), except for those that are known risk factors for sPTB/PPROM: a higher proportion of high-risk women with a recurrence, compared to those who delivered at term, had two or more previous early sPTB/PPROM events (31.8% vs. 9.2%) and multiple previous LLETZ or knife cone biopsies (13.6% vs. 1.1%). The median gestational age when the first valid VMB sample was taken was slightly later in the low-risk group (16^+5^ weeks) than in the high-risk group (16^+3^ weeks).

### VMB compositions and vaginal bacterial loads in high- and low-risk pregnant women

The VMBs of over 70% of pregnant women, high- and low-risk, were dominated by lactobacilli (defined as ≥75% relative abundance; Tables 2 and A3, heatmap in Figure A1). *L. iners* domination was most common overall (VMB type Li; 33.0% of high-risk and 22.1% of low-risk pregnant women), followed by *L. crispatus* domination (VMB type Lcr; 21.1% and 26.9%), and either domination by other lactobacilli (most commonly *L. gasseri* or *L. jensenii*) or ≥50% bifidobacteria (VMB type Lo+BL; 20.1% and 25.5%). In addition, 12.4% of high-risk and 12.8% of low-risk women had mild anaerobic dysbiosis (VMB type LA; defined as 25%-75% lactobacilli with the remainder BV-anaerobes), and 12.8% of high-risk and 13.1% of low-risk women had severe anaerobic dysbiosis (VMB type BV; ≥75% BV-anaerobes). Pathobionts and ‘other bacteria’ were rarely present, and if present, only in low relative abundance or estimated concentration (Tables 2 and A3).

**Table 2:**
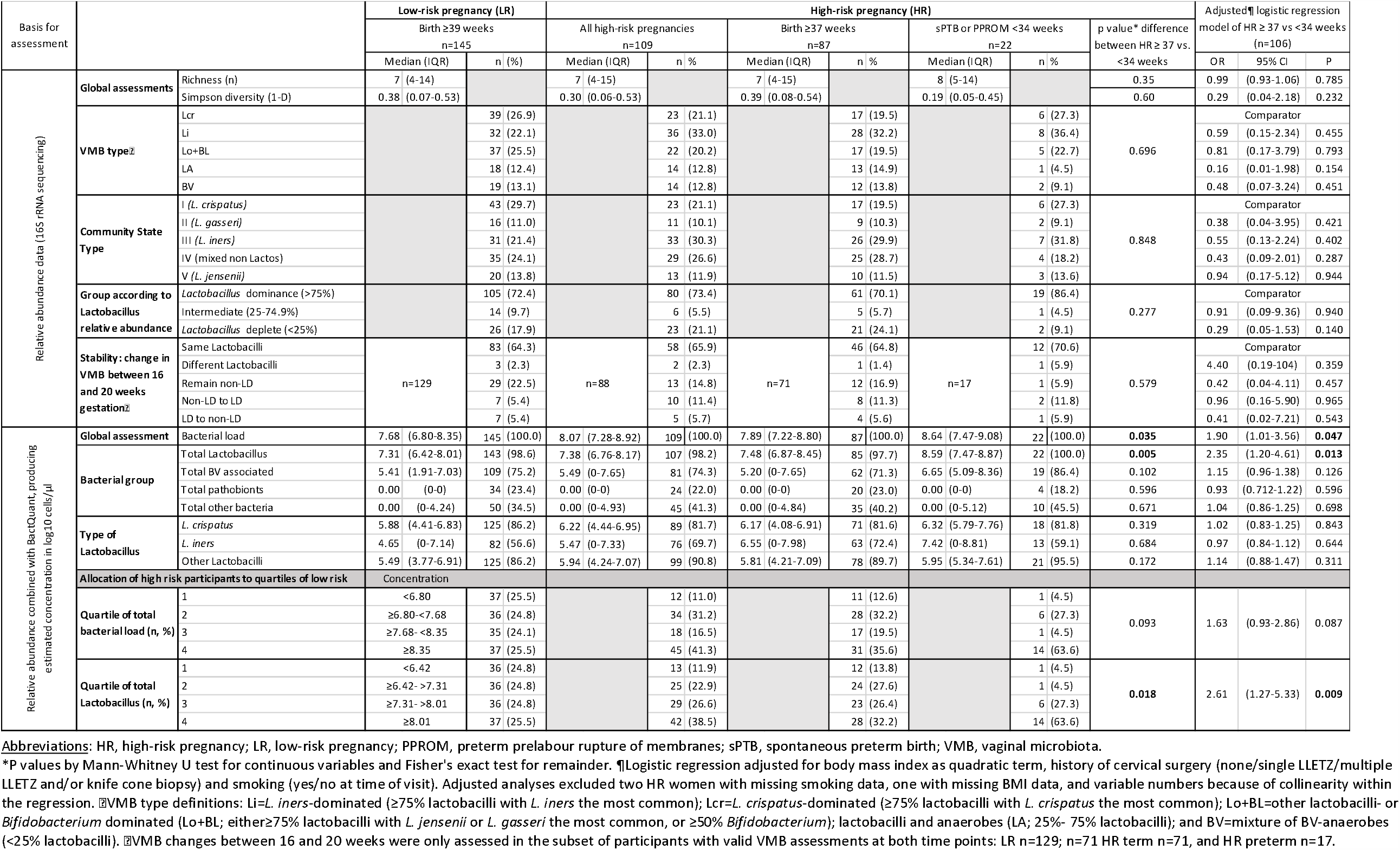
Associations between VMB variables and recurrent early sPTB/PPROM in high-risk women.

The VMB of the majority of participants with samples available from both visits between 15^+0^ and 23^+0^ weeks gestation was stable (Table 2). About two thirds of the high-risk (65.9%) and low-risk women (64.3%) were dominated by the same *Lactobacillus* species at both visits, and 14.8% of high-risk and 22.5% of low-risk women continued to have some degree of anaerobic dysbiosis. The remaining women switched *Lactobacillus* species (n=5) or shifted from lactobacilli-domination to anaerobic dysbiosis (n=12) or vice-versa (n=17).

The median vaginal bacterial load in the low-risk reference cohort was 7.68 log_10_ cells/μl (interquartile range (IQR) 6.80-8.35) and in the high-risk cohort 8.07 (IQR 7.28-8.92) log_10_ cells/μl (Table 2). High-risk women with mild or severe anaerobic dysbiosis (pooled together) had higher median vaginal bacterial loads than women with domination by *L. iners, L. crispatus*, or other lactobacilli/bifidobacteria (8.54, 7.79, 7.63, 7.53 log_10_ cells/μl, respectively; p-values are <0.05 for all comparisons between dysbiosis and the other groups, but are not significant for the comparisons between *L. iners*-domination and the other two lactobacilli groups; Table A4).

### Associations with early sPTB/PPROM recurrence – existing approaches

In the high-risk women, we did not identify any associations between recurrence and VMB richness, Simpson (1-D) diversity, CSTs,^15^ categorical *Lactobacillus* groups,^16^ categorical stability groups^17^ (Table 2), and presence or relative abundance of common *Lactobacillus* species (Table A3). *Ureaplasma* species and BVAB TM7-H1 were present in higher proportions of high-risk women with a recurrence than in high-risk women with a term birth, and *Dialister* species vice versa, but their relative abundances when present were very low (Table A3). In contrast, *Bifidobacterium breve* was present in a lower proportion of women with a recurrence, but its relative abundance when present was also low. The proportions of low-risk women with these taxa present were similar to those in high-risk women without a recurrence (Table A3, Figure A2A).

### Associations with early sPTB/PPROM recurrence – new approaches

We did not identify any associations between recurrence and VMB type or estimated BV-anaerobes or pathobionts concentrations (Table 2). However, high-risk women with a recurrence had a higher overall vaginal bacterial load (8.64 vs. 7.89 log_10_ cells/μl, adjusted odds ratio (aOR) 1.90, 95% confidence interval (CI) 1.01-3.56, p=0.047) and a higher total estimated *Lactobacillus* concentration (8.59 vs. 7.48 log_10_ cells/μl, aOR 2.35, 95% CI 1.20-4.61, p=0.013) than high-risk women without a recurrence (Table 2). The aOR was 1.63 (95% CI 0.93-2.86, p=0.093) for each increase in total vaginal bacterial load quartile, and 2.61 (95% CI 1.27-5.33, p=<0.001) for each estimated *Lactobacillus* concentration quartile (Table 2).

Table 3 shows estimated concentrations of total vaginal bacteria, the four bacterial groups, *L. cripatus, L. iners*, and other lactobacilli in high-risk women with and without a recurrence after stratification by VMB type. A trend towards higher recurrence risk with higher median bacterial loads was seen for each VMB type after stratification but was only significant in women with *L. iners*-domination: the median vaginal bacterial loads were 9.03 (IQR 8.02-9.12) log_10_ cells/μl in women with a recurrence compared to 7.79 (IQR 6.93-8.65) log_10_ cells/μl in women with a term birth (aOR 3.44, 95%CI 1.06-11.15, p=0.011). The association was no longer significant after adjustment for confounders (aOR 2.38, 95%CI 0.71-7.95, p=0.160). Within each of the other VMB type strata, the median estimated concentrations in high-risk women who had a term birth were similar to those in low-risk women, and non-significantly lower than those in high-risk women who had a recurrence.

**Table 3:**
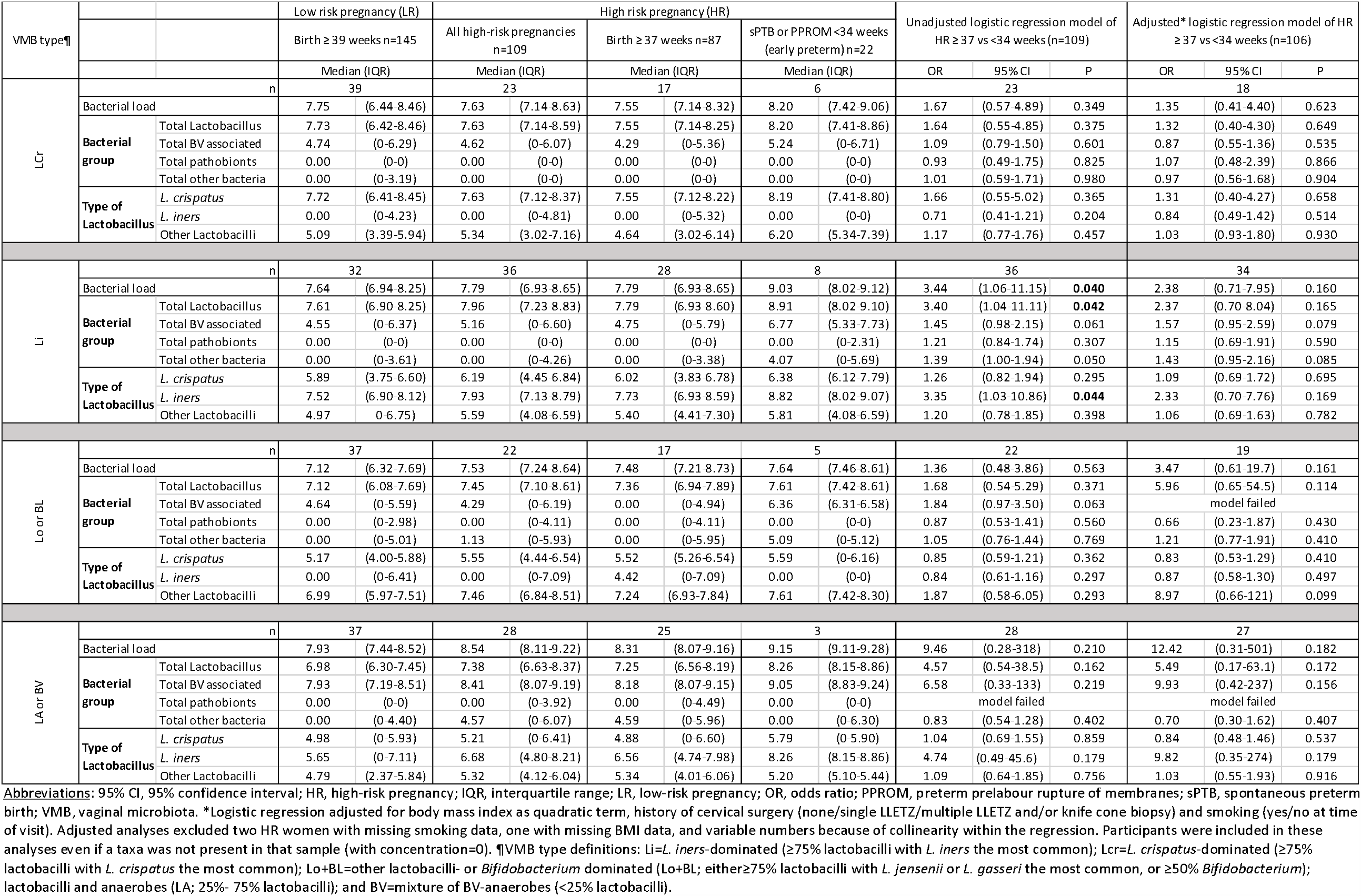
Associations between estimated median concentrations of vaginal bacteria and early recurrent sPTB/PPROM stratified by VMB type.

The majority of high-risk participants with an early sPTB/PPROM recurrence (17/22; 77.3%), and about half of the high-risk participants without a recurrence (44/87; 50.6%), had at least one VMB characteristic that was positively associated with recurrence. The following positively associated VMB characteristics overlapped considerably in both high-risk groups (Venn diagrams in Figure A2): highest quartile of total bacterial load, highest quartile of estimated total *Lactobacillus* concentration, highest quartile of estimated *L. iners* concentration, and presence of BVAB TM7-H1. The only positively associated VMB characteristic that did not completely overlap with the highest quartile of bacterial and estimated total *Lactobacillus* concentration was the presence of *Ureaplasma* species (this was the case in high-risk pregnancies with and without a recurrence).

## Discussion

### Main Findings

This exploratory study suggests that high vaginal bacterial load in the second trimester is associated with early sPTB/PPROM recurrence in high-risk pregnant women, irrespective of the bacterial composition. Our statistical power was limited: only 22 women had an early sPTB/PPROM recurrence. Therefore, not all associations in line with this conclusion reached statistical significance, especially after reducing statistical power further by stratification. However, we believe that this new hypothesis warrants further research for various reasons. First, the recurrence risk increased for each increase in bacterial and *Lactobacillus* concentration quartile (on a log_10_ scale). Second, the findings were consistent for each VMB type after stratification: higher recurrence in high-risk women with higher median bacterial loads for all VMB types (reaching significance for *L. iners*-domination), and similar median bacterial loads in high-risk and low-risk women who had a term birth for all VMB types. Third, estimated concentrations reached higher levels for anaerobic dysbiosis and *L. iners*-domination than for domination by other lactobacilli or bifidobacteria. Our findings therefore do not contradict earlier findings by other groups that indicated increased PTB risk for anaerobic dysbiosis and high *L. iners* relative abundance.^8,22–24^ In fact, the inconsistent findings related to *L. iners* in the literature might be due to the lack of quantification in most studies.

### Interpretation in light of other evidence

To our knowledge, the potential association between the vaginal bacterial load and PTB has only been assessed in two previous studies. Freitas et al^7^ also showed a higher 16S gene concentration in the second trimester (approximately 16 weeks gestation) in women who had an sPTB <37 weeks compared to women with a term birth ≥37 weeks (8.1 vs. 7.8 log_10_ copies/swab, respectively). Conversely, Elovitz et al^8^ reported a similar 16S gene concentration in women who had a sPTB <37 weeks compared to women with a term birth ≥37 weeks in samples collected before 24 weeks gestation (approximately 8 log_10_ copies/swab in each group), and a lower concentration (approximately 7.5 vs 7.9 log_10_ copies/swab) in samples collected at 28 weeks gestation. High vaginal bacterial load may only be associated with PTB when present prior to 24 weeks gestation, but additional quantitative studies are needed.

Female genital tract inflammation is considered one potential cause of PTB.^25^ BV (a clinical condition caused by anaerobic dysbiosis) is associated with vaginal inflammation, and is a very common condition in non-pregnant women.^26^ Pregnant women are protected from BV due to the high oestrogen levels during pregnancy.^27,28^ In our study, the prevalence of anaerobic dysbiosis, and the relative abundances and estimated concentrations of BV-associated bacteria and pathobionts, were indeed low. This likely explains why we did not identify any statistically significant associations between the presence or extent of anaerobic dysbiosis and early sPTB/PPROM recurrence. However, the non-significant trends consistently suggested increased risk. Another common inflammatory vaginal condition is vulvovaginal candidiasis, for which pregnant women are at increased risk.^29^ In our study, only one woman (low-risk) reported having been treated for vulvovaginal candidiasis prior to study enrolment. Future studies should systematically assess vaginal yeasts concentrations in addition to bacterial concentrations in all participants, and determine their potential contributions to early sPTB/PPROM recurrence.

In our study, *L. iners* was able to achieve higher bacterial loads than the other lactobacilli, and was associated with early sPTB/PPROM recurrence. *L. iners* has a genome of just 1.3 Mbp,^30^ strikingly smaller than the other common vaginal *Lactobacillus* species, and seems fully adapted to the vaginal niche.^31^ This may explain why it is able to grow to such high densities in the vagina. *Lactobacillus* species are considered to be non-inflammatory when present in the vagina,^32^ but they might cause inflammation when they travel through the cervical mucus plug into the upper genital tract. Ascension to the upper genital tract may be increased when vaginal density is high. Studies have shown that *L. iners* with high expression of clustered regularly inter-spaced short palindromic repeat (CRISPR) genes are present in women with BV but not in women without BV.^33^ CRISPR genes are the primary bacterial defence against bacteriophages. The *L. iners* strains associated with high bacterial loads and recurrent sPTB/PPROM in our study may differ from strains associated with lower bacterial loads. This requires further research.

Two uncommon taxa (present in fewer than 20% of the women in the low risk group) were non-significantly associated with early sPTB/PPROM recurrence in our study and were also associated with PTB in other studies: *Ureaplasma*^7,24^ and BVAB TM7-H1^6^. In women with early sPTB/PPROM recurrence, the presence of *Ureaplasma* species in the vagina did not completely overlap with having high bacterial loads and lactobacilli concentrations. *Ureaplasma* species may therefore play a role in PTB that is independent from overall vaginal bacterial composition and load. Our data confirms that *Bifidobacterium breve* may have a protective effect against early sPTB/PPROM recurrence.^16^

### Strengths and Limitations

Our study had some limitations, most notably limited statistical power and insufficient data on vaginal yeasts, but also had strengths. Pregnancy outcomes were very carefully selected and assessed, many VMB composition variables were created covering different aspects of VMB composition, and most of these VMB variables were (semi)quantified. The latter turned out to be crucial, as our most important findings concern bacterial concentrations. The quantification method that we used has recently been validated for non-minority taxa in two different studies.^7,8^ We had initially planned to analyse early sPTB and early PPROM as two separate outcomes because the aetiological pathways may differ,^34^ and to exclude women who used PTB prevention treatments,^35^ but the limited statistical power did not permit this. A final limitation is the lack of VMB data in the third trimester and/or closer to the birth.

## Conclusion

Vaginal bacterial load in the second trimester, irrespective of the bacterial composition, was associated with early sPTB/PPROM recurrence. Domination by lactobacilli other than *L. iners* may protect women from developing high bacterial loads. These findings should be confirmed in larger, longitudinal studies that incorporate quantification of vaginal bacteria and yeasts. If they are confirmed, interventions that maintain a non-*iners* lactobacilli/ bifidobacteria-dominated VMB may protect women from inflammation-associated PTB.

## Data Availability

Once accepted for publication in a journal the authors agree to share the data on http://datacat.liverpool.ac.uk/view/.

## Acknowledgements

We would like to thank all participants for their enthusiastic involvement in the study, in particular members of the Harris-Wellbeing Patient and Public Engagement group. We would also like to thank the Liverpool Women’s Hospital for hosting the research, Tracy Ricketts at the Liverpool Women’s Hospital for administrative support, University of Liverpool Centre for Genomic Research staff for 16S sequencing, and Jacques Ravel and Mike Humphrys at the University of Maryland School of Medicine, Institute for Genome Sciences, for the 16S qPCR assays.

## Contribution to authorship

JW, AS, DR, BM-M, AC, AA and ZA conceived the study, wrote the protocol and obtained funding. AC, JI, BP and LG contributed to the protocol and recruited participants. AD, JW, aCG, and CB developed the DNA extraction, sequencing, and bioinformatics methods. LG, AC and AS extracted the clinical data. LG performed the laboratory analysis. JW developed the VMB analysis methods, and LG and MV performed the data processing. LG performed the data analysis and wrote the initial draft, and JW, ZA and MV contributed to data interpretation and revised the paper. All authors approved the final manuscript.

## Details of Ethics Approval

The study was approved by North West Research Ethics Committee - Liverpool Central, reference 11/NW/0720 on 4 November 2011. The collection of vaginal fluid for this component of the study was approved in amendment 4 on 19 October 2015.

## Tables, Figures, and Supplementary Materials Caption List

Appendix A: Supplementary methods, figures, and tables

Appendix B: Assignment of amplicon sequence variants to bacterial groups

