## Appendix A for "Vaginal bacterial load in the second trimester is associated with early preterm birth recurrence: a nested case-control study"

### Supplementary methods

We conducted an observational study to assess the association between the vaginal microbiota (VMB) and recurrent early preterm birth. This was a sub-study within the “Development of novel biomarkers for prediction of preterm labour in a high-risk population” study. The study was funded by a charitable donation from Lord and Lady Harris, gained in an open competition that was facilitated by the charity Wellbeing of Women. The study was approved by the North West Research Ethics Committee - Liverpool Central, reference 11/NW/0720, on 4 November 2011. Recruitment started on 1 April 2012. The VMB sub-study was added on 19 October 2015 via protocol amendment. Recruitment was completed on 31 December 2017. All participants gave written informed consent.

#### Eligibility criteria

We recruited two cohorts of pregnant women: a high-risk cohort and a low-risk cohort.

The eligibility criteria for both cohorts were:

- Singleton pregnancy.
- Having had an early pregnancy ‘dating’ scan^1^ before 14^+0^ weeks gestation showing no foetal abnormalities. This scan was used to calculate gestational age at sampling and birth.
- Between 15^+1^ and 23^+0^ weeks gestation at recruitment.
- Having taken part in standard antenatal care, including HIV and hepatitis B testing at the booking appointment (first appointment with a midwife).^1^ The results of these tests had to be negative for blood handling safety reasons. The uptake of HIV testing in UK antenatal services in 2015 was 98.2%, with only 0.15% of women testing positive,^2^ so this is unlikely to have had a substantial impact upon recruitment.
- Not having had vaginal sex within 48 hours prior to the first study visit. All women who had a second study visit were asked to abstain from sex for 48 hours prior to the study visit.
- Participants were only eligible for study participation once and could not be recruited a second time during a subsequent pregnancy.

Additional eligibility criteria for the high-risk cohort were:

- Having had at least one previous spontaneous preterm birth (sPTB), preterm prelabour rupture of membranes (PPROM), or spontaneous late miscarriage at 16^+0^-33^+6^ weeks gestation.
- Not using preterm birth preventative treatment (cervical cerclage, vaginal progesterone, or a vaginal pessary) at the time of recruitment.
- Women were not excluded if they had had significant cervical surgery or previous obstetric or medical problems.

Additional eligibility criteria for the low-risk cohort were:

- Having had at least one previous pregnancy that progressed beyond 37^+0^ weeks gestation.
- No history of late miscarriage (beyond 16^+0^ weeks gestation).
- No history of preterm birth (<37^+0^ weeks gestation), including medically induced preterm birth.
- No history of PPROM (<37^+0^ weeks gestation).
- No history of significant cervical surgery, defined as either: 1 large loop excision of the transformation zone (LLETZ) over 10mm; 2 or more LLETZ; or a knife cone biopsy.
- No medical condition considerably affecting health, defined as having been referred to a specialist antenatal clinic for a medical condition by the booking midwife. This includes, but is not limited to, epilepsy, inflammatory bowel disease, or renal impairment.
- No previous pregnancy affected by an obstetric complication with an increased chance of reoccurrence, defined as previous gestational diabetes, preeclampsia requiring magnesium sulphate, or gestational hypertension requiring antihypertensive medication.

#### Rationale for inclusion of the low-risk cohort

The low-risk cohort was designed to reflect the ‘healthy’ pregnancy population as much as possible. Participation was limited to parous women only because the high-risk cohort consisted of parous women by definition. VMB compositions are known to be influenced by local factors such as the age and ethnicity distribution, lifestyle factors (including diet and hormonal contraception use), and environmental factors.^3,4,5,6^ We wanted to characterise the expected distribution of VMB parameters in our local population because this had never been done before. In addition, there is no consensus yet on how best to analyse VMB sequencing data.^7,8^ The low-risk cohort allowed us to describe the expected distribution of VMB parameters using our own analysis methods.

#### Recruitment procedures

In the UK, all pregnant women are encouraged to book with an antenatal care provider in the first trimester, and the majority of women will continue to receive antenatal and intrapartum care from that same provider.^1^ Liverpool Women’s Hospital^9^ is one such care provider, with the largest stand-alone maternity unit in the UK and a dedicated midwifery led birthing suite. The hospital provides ‘low-risk’ pregnancy care, including ‘midwifery led care’,^1^ as well as high-risk pregnancy care. High-risk women were recruited from the Liverpool Women’s Hospital Preterm Labour Prevention Clinic. Low-risk women were recruited from the regular antenatal care clinic at Liverpool Women’s Hospital; they planned to either give birth at Liverpool Women’s Hospital or with the hospital’s linked homebirth team.

Low risk participants were recruited as follows:

1. Study clinicians used hospital electronic records to identify women who had booked for antenatal care^1^ at Liverpool Women’s Hospital and fitted the eligibility criteria as described above. Study staff then phoned or sent a text message to potential participants between the early pregnancy dating scan (described above) and 15^+0^ weeks gestation to offer them study participation. Each potential participant was only contacted once, and a voicemail was left if the phone was not answered and voicemail was available.
2. Low-risk women were also approached by study leaflet (distributed by community midwives) or social media (distributed by research staff or the Liverpool Women’s Hospital communications team). Potential participants were invited to phone or send a text message to a dedicated telephone number.
3. Study staff spoke to potential participants on the telephone. During this conversation, they confirmed eligibility and explained the study, including the rationale and need for a speculum examination with vaginal swabbing and a cervical length scan. Participants were offered a 3D ultrasound scan of the pregnancy at each study visit.

High risk participants were recruited as follows:

1. Women with at least one sPTB, PPROM, or spontaneous late miscarriage at 16^+0^-33^+6^ weeks gestation were routinely referred to the hospital’s Preterm Birth prevention Clinic by their booking midwife. This was part of standard antenatal care for all eligible patients.
2. Study clinicians reviewed the patients’ clinical notes to confirm eligibility (primarily not using preterm birth prevention therapy at the time of study recruitment).
3. Study clinicians then approached potential participants during their preterm birth prevention clinic visits to offer study involvement.

These procedures were designed with input from the Liverpool Women’s Hospital patient and public involvement group (see below), and were approved by the ethics committee. The use of electronic case notes for participant identification predated changes to UK general data protection legislation.^10^

#### Clinical procedures

Study visits of high-risk women were timed to coincide with the participants’ scheduled visits to the Liverpool Women’s Hospital Preterm Labour Prevention Clinic, and took place at approximately 16 weeks (15^+1^-18^+6^ weeks) and 20 weeks (19^+0^-23^+0^) gestation. Low-risk women also had scheduled study visits at approximately 16 weeks (15^+1^-18^+6^ weeks) and 20 weeks (19^+0^-23^+0^ weeks) gestation. In both cohort, the second study visit was performed a minimum of two weeks after the first study visit. Participants who were recruited after 19^+0^ weeks gestation only had a single study visit.

At their first visit, an obstetrician confirmed eligibility and gave the potential participant an information leaflet to read. Women were given the opportunity to ask questions and written informed consent was obtained. If a woman was found to be ineligible or declined to participate, the 3D ultrasound scan was still performed.

Next, participants underwent the following procedures by an obstetrician:

1. A face-to-face interview about demographic factors, height, weight, smoking, antimicrobial use during pregnancy, and medical and obstetric history.
2. Blood sampling (for another part of the project; results not described in this paper).
3. A speculum examination with a chaperone present. During this examination, cervicovaginal fluid was taken from the posterior fornix (10 seconds of rotation) for quantitative foetal fibronectin (qfFN) assessment; three high vaginal swabs using HydraFlock standard tapered swabs (Medical Wire and Equipment, Corsham, England) for VMB assessments; and a high vaginal charcoal swab for routine processing by the local NHS laboratory.
4. A cervical length scan.
5. 3D ultrasound scan. Photographs were provided to the participant.

Any questions were addressed as necessary and the second clinic visit was scheduled.

Samples were processed on the same day. The qfFN swab was tested on a Rapid fFN 10Q System (HOLOGIC, Marlborough, MA, USA), but the study obstetricians were blinded to the qfFN result until all participants in the study had given birth. One HydraFlock swab was placed in 1ml of RNAlater (Merck Life Science UK Ltd, Dorset, UK), and the other two were stored dry, in 1.2ml cryogenic tubes (Fisher scientific, Loughborough, UK) at -80°C within an hour of sampling. The charcoal swab was transported to the local NHS laboratory for diagnosis of bacterial vaginosis and candidiasis by culture. The latter results were not available to the study team, but only one woman reported having received treatment for vulvovaginal candidiasis during her pregnancy, prior to enrolment into the study.

At the second study visit, an obstetrician verbally confirmed ongoing willingness to participate in the study and questioned the participant about any new conditions that had been identified during the pregnancy, use of preterm birth prevention treatment, antimicrobial medication use, any other medication use in the previous 72 hours (including vitamins), and vaginal sex in the previous 48 hours. The blood sampling, speculum examination with vaginal swabs, cervical length scan, and 3D ultrasound scan were then repeated in the same way as described above for the first visit. At the end of the second visit, contact details were confirmed, and participants were asked for their permission to be contacted about the pregnancy outcome should the need arise (see below).

Pregnancies were managed in accordance with usual practices in the Liverpool Women’s Hospital. Preterm birth prevention therapy (cerclage, vaginal pessary, or vaginal progesterone) was offered if cervical length was ≤ 25mm or based on clinician and patient preference in case of a large change in cervical length between measurements. None of the low-risk women required an intervention but most (78/109) of the high-risk women did.

We were not able to collect data about antimicrobial use in pregnancy – other than by participant self-report - because of the wide range of treatment providers. These include the hospital, primary care practitioners, walk-in centres, or - in the case of vulvovaginal candidiasis - over the counter purchase.

#### Ascertainment of outcomes and treatments after pregnancy

Liverpool Women’s Hospital records were reviewed by study obstetricians to ascertain pregnancy outcomes and whether any preterm birth prevention treatment had been used. Birth outcomes were classified independently by two clinicians experienced in PTB management. When there was a discrepancy between them, the case was reviewed by a third experienced clinician until the team reached consensus. If a participant eventually delivered elsewhere, pregnancy outcome information was requested from the participant’s delivery care provider, or (if this was not successful) from the participant herself.

#### Laboratory procedures

*DNA extraction*

DNA extraction and sequencing were done at the University of Liverpool Centre for Genomic Research. DNA was extracted from one sample per participant per visit. A total of 706 swabs from 364 participants underwent DNA extraction, PCR amplification, and 16S rRNA sequencing. Any subsequent data processing was restricted to data from the 259 participants that were included in the high- and low-risk cohorts of the case-control sub-study.

The samples were thawed, and DNA was extracted by adding 180 μl of enzymatic lysis buffer containing lysozyme to the sample (Sigma-Aldrich, Dorset, UK); incubation for 30 minutes at 37 °C; adding 25 μl of proteinase K and 200 μl of buffer AL from the Qiagen DNeasy Blood and Tissue kit (Qiagen, Manchester, UK); incubation for 30 minutes at 560 C; adding 200 mg of 0.1 mm zirconia/ silica beads (Thistle Scientific, Glasgow, UK), and bead-beating for 5 minutes at 25 Hz on a Qiagen TissueLyser II (Qiagen, Manchester, UK). Next 200 μl of 100% ethanol was added to the sample followed by centrifugation. The swab head was discarded, and the pellet was purified in four subsequent centrifugation steps after adding one-by-one 200 μl 100% ethanol, 500 μl buffer AW1, 500 μl buffer AW2 and 75 μl buffer AE as per manufacturer’s instructions (Qiagen, Manchester, UK). In order to facilitate detection of contaminants downstream, we included one negative control (an empty tube) with each DNA extraction round of 23 study samples. The DNA concentration of all samples was measured by Qubit (Invitrogen, Thermo Scientific, Paisley, UK) and the DNA quality of all samples by Nanodrop (Thermo Scientific, Paisley, UK). Samples with a particularly low DNA concentration or quality were discarded (n=25) and the DNA extraction process was performed on the second sample taken during that study visit (these samples subsequently achieved acceptable concentration and quality assessments).

*PCR amplification and 16S rRNA gene sequencing*

Two PCR rounds were performed for each DNA sample (study samples and negative controls) for 16S rRNA gene amplification and barcoding. Firstly, the V3-V4 region of the 16S rRNA gene was amplified as described previously.^11^ DNA was amplified using 1.25 μl of a 10 µM concentration of 319F forward primer (5’-ACTCCTACGGGAGGCAGCAG-3’) and 1.25 μl of a 10 µM concentration of 806R reverse primer (5’-GGACTACHVGGGTWTCTAAT-3’), 12.5 μl NEB Next HF 2x PCR Master Mix (New England Biolabs, Hitchin, UK), 9 μl of nuclease-free water and 1 μl of DNA extraction product to make a 25 μl reaction volume. The first denaturation cycle was performed for 30 seconds at 98 °C, followed by 10 cycles with a denaturation cycle of 10 seconds (at 98 °C), an annealing cycle of 30 seconds (at 58 °C), an extension cycle of 30 seconds (at 72 °C), and finally an extension cycle of 5 minutes at 72 °C. PCR products were then purified and size-selected using Agencourt AMPure XP beads (Beckman Coulter, High Wycombe, UK) in a 1:1 bead-to-sample ratio. The final PCR round used the standard Illumina Nextera XT index kit v2 (Illumina, San Diego, CA, USA), aimed at V3-V4 sequences by a dual-index approach. This permits multiplexing of up to 384 samples at a time (two rounds were performed to accommodate all samples). The barcoding used 2.5 μl of Index 1 primer, 2.5 μl of Index 2 primer, 12.5 μl NEB Next HF 2x PCR Master Mix and 7.5 μl sample making a 25 μl reaction volume. The first denaturation cycle was performed for 3 minutes at 98 °C, followed by 15 cycles with a denaturation cycle of 30 seconds (at 98 °C), an annealing cycle of 30 seconds (at 55 °C), an extension cycle of 30 seconds (at 72 °C), and a final extension cycle of 5 minutes at 72 °C. AMPure beads were then used to purify PCR products as explained above, also using a 1:1 bead-to-sample ratio. Each PCR run also had a negative control (10 μl of nuclease-free water instead of 9 μl of nuclease-free water and 1 μl of DNA) to identify contaminants, and 10 μl of 0.2 ng/μl ZymoBiomics Microbial Community DNA standard (Zymo Research Corp, Irvine, CA, USA), a commercially available positive control. The PCR runs also included the DNA extraction negative controls. DNA collected from the same participant at different visits were included in the same PCR runs. The Qubit Fluorometer with the dsDNA HS Assay kit (Invitrogen, Thermo Scientific, Paisley, UK) was used to measure PCR product DNA concentrations of each sample (including negative and positive controls). Two negative controls were not successfully amplified and were excluded from the subsequent steps; all participant samples, positive controls, and the remainder of the negative controls were used.

Amplicons from samples were evenly pooled into sequencing libraries at a mass of 0.8 ng DNA per amplicon. To achieve this, Qubit DNA concentrations and Fragment Analyzer (Agilent, Santa Clara, USA) quality control information were combined for pooling before size-selection using Pippin Prep (Sage Scientific, Beverly, Massachusetts, USA). Samples with a DNA concentration of <0.30 ng/µl (such as the negative controls) were added in a fixed volume of 1 µl. The two libraries were sequenced on an Illumina HiSeq instrument (Illumina, San Diego, CA, USA), run in rapid mode, 2x300bp using a 250PE and 50PE kit. DNA collected from the same participant at different visits was included in the same library.

*Panbacterial 16S rRNA gene qPCR*

Extracted DNA from all participant samples (706 samples from 364 participants) was sent to the Institute for Genome Sciences of the University of Maryland (Baltimore, MD, USA) for estimation of the panbacterial 16S rRNA gene copy concentration using the BactQuant qPCR assay. This assay is based on an analyses of 4,938 16S rRNA gene sequences in the Greengenes database.^9,10^ The analysis was performed as described previously.^10,11^ Briefly, 1.5 μl of template (1:10 diluted DNA) was added to 3.5 μl of reaction mix, with the final reaction containing 1.8 μM each of the forward (341F) and reverse (806R) primer targeting the 16S V3-V4 region, 225 nM of the TaqManW probe, 1x Platinum Quantitative PCR SuperMix-UDG with ROX (Invitrogen, Thermo Scientific, Waltham, MA, USA) and molecular-grade water. Each sample, including no template controls, were assayed in triplicate. An in-run standard curve (ranging from 10 to 10^8^, with 10^2^–10^8^ in 10-fold serial linear dilutions) was used in each run. Amplification and real-time fluorescence detections were performed on the Bio-Rad CFX 384 instrument (Bio-Rad Inc., Hercules, CA, USA). The PCR conditions were: 3 minutes at 50 °C for UDG treatment, 10 minutes at 95 °C for Taq activation, 15 seconds at 95 °C for denaturation and 1 minute at 60 °C for annealing and extension, times 40 cycles. Cycle threshold (Ct) value for each 16S qPCR reaction were obtained using a manual Ct threshold of 0.05 and automatic baseline. The 16S rRNA gene concentration was reported in copies/μL for each sample.

Quality control of the BactQuant assay was performed by excluding samples that did not amplify in two of three, or all three, of the triplicate assays, or had skewed low 16S rRNA gene concentration results of <1,000 copies/μl. This was the case for 31 of 706 samples.

#### Molecular data processing

*Sequencing data processing*

The mean raw unpaired read count of all 706 samples from 364 participants was 368,205 reads per sample (95% confidence interval (CI) 353,388 – 383,022 reads). Cutadapt v1.16^12^ was used to demultiplex the reads and remove primer sequences. All subsequent steps were performed in DADA2 version 1.8 package for large paired-end datasets in R version 3.5.1 (R core team, 2015).^13^ DADA2 was chosen because of its ability to resolve reads to a single nucleotide. The fastqFilter command was used for error correction with parameter settings aiming to maximize read retention. The minimum read lengths (truncLen) were set to 255 for forward reads and 210 for reverse reads based on the quality plots, maxEE to a maximum of 5 for forward read and 8 for reverse read expected errors, maxN to zero ambiguous bases allowed, and truncQ to zero. Approximately 10% of reads were discarded after error correction. The learnErrors command was then used to determine the read error rates. The reads were assigned to unique amplicon sequence variants (ASVs, equivalent to a taxon) using the derepFastq command, and ASVs with higher than average error rates were discarded (denoised) using the dada command.^13,14^ The mergePairs command was used to merge forward and reverse reads. The removeBimeraDenovo command was used to remove chimeric compositions of two separate parent ASVs (Bimeras) with the Silva version 132 database as the reference database;^15^ 6.3% of ASVs were identified as bimeric and removed. Overall, a median of 28% of the raw reads per sample were removed during these DADA2 clean-up processes.

DADA2 was used to perform taxonomic assignment in two steps. Firstly, assignTaxonomy was used to map ASVs to taxa at genus level or above using the RDP classifier with a minimum bootstrap value of 50% and the Silva v132 database as the reference database.^15,16^ Secondly, addSpecies was used to map ASVs to species level. Only ASVs with exact (100%) identity matches with species in the Silva database were assigned to each species.

Next, a spreadsheet containing the sequences, taxonomic assignments, and read counts for each ASV per sample was imported into Microsoft Excel for Mac version 16. ASVs with a read count in all samples combined of less than 100 were removed, as well as two non-bacterial ASVs, and one likely contaminant ASV. The likely contaminant was a *Cutibacterium* genus that was present in two negative controls at a relative abundance that was higher than in any study sample. The Silva v132 database did not include the vaginal taxa BV-associated bacterium 1 (BVAB1), BVAB2 and BVAB TM7 sequences. These have, however, been published elsewhere^17,18^ and were manually identified in our dataset. We also searched for *Mageeibacillus indolicus* (BVAB3) and *Fenollaria massiliensis* sequences manually, but these were not identified. The taxonomic assignments derived from the Silva database were manually double-checked for the 112 ASVs with a relative abundance of at least 0.05% of the read count of all samples combined (out of a total of 1646 ASVs), using the Microbial Nucleotide BLAST (BLASTn) function on the National Center for Biotechnology Information NCBI website.^19^ In cases of a discrepancy, the Vaginal 16S rDNA Reference Database was used as a tiebreaker.^20^ This resulted in 45 *Lactobacillus* genus ASVs being reassigned to various *Lactobacillus* species, two *Streptococcus* genus ASVs being reassigned to *S. agalactiae*, a *Staphylococcus* genus being reassigned to *S. aureus*, a *Gardnerella* genus ASV being reassigned to *G. vaginalis*, an *Atopobium* genus ASV being reassigned to *A. vaginae*, a *Sneathia* genus ASV being reassigned to *S. amnii* and an *Enterococcus* genus ASV being reassigned to *E. faecalis*. Read counts for ASVs assigned to the exact same taxonomy were then summed for each sample. The lowest total read count for any specific sample above 1000 reads was 1101, so GuniFrac 1.1 package in R was used to rarefy to 1101 reads. The rarefied ASV table contained 690 (of the original 706) samples and 290 unique ASVs. The prop.table function in R was then used to transform rarefied read counts into relative abundances.

*Estimation of bacterial load and taxa concentration*

The overall bacterial load and ASV-specific concentrations per sample were estimated by combining the sample-specific 16S rRNA gene concentration data with the rarefied ASV table. The 16S rDNA gene copy number was identified for each of the 276 unique ASVs in the final rarefied ASV relative abundance table using the NCBI version of the rrnDB database,^21^ or the Greengenes database in case of missing data.^9^ In situations where ASVs were mapped to multiple species at genus level, the mean 16S gene copy number based on all potential species was used. When the mean 16S gene copy number of a species was not available, the mean copy number of the corresponding genus was used. For BVAB1 and BVAB2 only ‘order’ level taxonomic information (Clostridiales order). We used the Clostridiales order mean copy number (=4.62) for BVAB1 and BVAB2 because data for lower level taxonomies were not available. The ASV-specific copy-normalized rarefied relative abundance was multiplied by the sample-specific 16S rRNA gene copies concentration to estimate the concentration of each ASV in cells/μl per sample. This method has previously been shown correlate with species-specific qPCR results for non-minority species.^22,23^ These concentrations were log_10_-transformed. To prevent skewed negative values, concentration results less than one cell/μl were set to one prior to log_10_-transformation.

#### Selection of participants for nested case-control study

Within the high-risk group, 109 of 133 enrolled women were retained for analysis: 87 participants who gave birth ≥37^+0^ weeks gestation, without PPROM, and 22 participants who had sPTB or PPROM <34^+0^ weeks gestation (main manuscript Figure 1). High-risk participants were retained if they received PTB prevention therapy after recruitment as long as the first sample available for analysis (i.e. a sample that passed quality control) was taken prior to using PTB prevention therapy. Iatrogenic preterm births (n=8) were excluded.

Within the low-risk group, 145 of 217 enrolled women were retained for analysis. All of them gave birth at ≥39^+0^ weeks gestation. We had planned to exclude women who received preterm birth prevention treatment, but none of the women required such treatment.

We only analysed low-risk women who gave birth ≥ 39^+0^ weeks gestation because previous work suggests that these births represent truly healthy term birth.^12^ Mortality and morbidity are higher amongst infants born at 37 to 38 weeks gestation, compared to those born at 39 to 40 weeks gestation.^13–16^

A total of 259 of the 364 participants had an eligible pregnancy outcome. These participants contributed 484 samples. Of these, 454 (93.8%) samples from 254 (98.1%) participants had valid 16S rRNA sequencing and valid BactQuant assays and were retained for further analysis. The final rarefied ASV relative abundance table consisted of 276 ASVs in 454 samples from 254 individual participants, mapping to species (181; 65.6%), genus (82; 29.7%), or higher taxonomic levels (13; 4.7%) (Appendix B). Of the 276 ASVs, 76 (27.5%) ASVs were present at a relative abundance of at least 1% in at least 1 sample; the other 200 (72.5%) ASVs were minority species.

#### Creation and selection of VMB variables

As explained in the manuscript, we use existing approaches of sequencing data reduction to create VMB variables, but we also used new approaches that have not been used in PTB research before.

*Existing approaches*

We systematically reviewed the published literature to identify VMB variables that were associated with PTB in at least one study (Table A1). Table A1 shows previous studies that used sequencing methods to determine the relationship between the VMB and PTB. The first author (LG) first extracted relevant information from the main manuscripts of each study to create a list of VMB variables that were described in at least one study. Next, she assessed each manuscript again for any data related to any of the variables on the full list of VMB variables, including data in supplementary materials. Individual taxa were often shown in heatmaps or other figures without information about a statistical relationship with PTB, or a relationship was inferred but not clearly described. Such cases are shown as grey cells in Table 1, either empty (no information) or containing a horizontal arrow (ambiguous information). Table A1 was double-checked by one of the senior authors (JW).

We selected variables that had been associated with PTB in at least two studies, irrespective of the directions of those associations (positive or negative). These included: richness and Simpson diversity (1-D) as continuous variables; two categorical VMB composition variables with each participant assigned to one category (the community state types (CSTs) described by Ravel et al^14^ and *Lactobacillus* groups (dominant, intermediate, deplete) based on *Lactobacillus* relative abundance;^15^ stability groups, slightly modified from Romero et al;^16^ and the presence/absence of 23 specific taxa of interest. We included some individual taxa of interest even though they had only been associated with PTB in a single study: BVAB1, BVAB2, and BVAB TM7 (because they had only recently been described at the time the reviewed studies were conducted), *L. jensenii* (because we wanted to include all four major vaginal *Lactobacillus* species), and *Bifidobacterium breve* (because of its potential role as a preventative probiotic).

We used hierarchical clustering analysis with ward linkage and a clustering density threshold of 0.75 using all available data to assign samples to one of five CSTs: I (*L. crispatus*-dominated), II (*L. gasseri*-dominated), III (*L. iners*-dominated), IV (mixed bacterial species), and V (*L. jensenii*-dominated). The three *Lactobacillus* groups were defined by *Lactobacillus* relative abundance as follows: dominant (≥75%), intermediate (25-74.9%), and deplete (≤25%). VMB stability was only assessed in the subset of participants who had valid VMB results available for both study visits: 17/22 (77.3%) high-risk participants with early sPTB/PPROM, 71/87 (81.6%) high-risk participants with term births, and 129/145 (89.0%) low-risk participants with term births. Based on a modified version of the stability groups described by Romero *et al*^17^ each participant was allocated to one of five stability groups: 1) same lactobacilli ( ≥75% lactobacilli (=lactobacilli-dominant or LD) at both visits with dominance of the same species); (2) different lactobacilli (LD at both visits with dominance of different species); (3) persistently non- LD (non-LD at both visits); 4) non-LD to LD (non-LD at the first visit and LD at the second visit); and 5) LD to non-LD (LD at the first visit and non-LD at the second visit).

*New approaches*

The final set of VMB variables was based on our previous work and was applied to PTB research for the first time: VMB types (based on relative abundances), bacterial group relative abundances, and bacterial group estimated concentrations. We also included total bacterial load, which had been considered in a few previous studies but not in much detail.^18,19^

The VMB types were mutually exclusive and were defined as follows: *L. iners*-dominated (Li; ≥75% lactobacilli of which *L. iners* was the most common); *L. crispatus*-dominated (Lcr; ≥75% lactobacilli of which *L. crispatus* was the most common); other lactobacilli-dominated or *Bifidobacterium*-dominated (Lo+BL; ≥75% lactobacilli of which *L. jensenii* or *L. gasseri* were the most common or ≥50% *Bifidobacterium*); a mixture of lactobacilli and anaerobes (LA; 25%-75% lactobacilli and the remainder BV-anaerobes); and a mixture of BV-anaerobes (BV; ≥75% BV-anaerobes).

Each non-minority amplicon sequence variant in each sample was allocated to one of four bacterial groups based on the published literature (Appendix B): lactobacilli; BV-anaerobes (consisting of Actinobacteria, Bacteroidetes, Firmicutes, Fusobacteria, and Tenericutes except those included in the other 3 groups); pathobionts (most Proteobacteria, and streptococci, staphylococci, enterococci, Spirochaetaceae, *Listeria*, *Chlamydia trachomatis*, and *Neisseria gonorrhoeae*); and other bacteria (a rest group, containing Actinobacteria that are known to be (facultative) aerobic skin bacteria, *Bifidobacterium* species, and difficult-to-classify minority species). Within each sample, read counts of ASVs belonging to the same bacterial group were summed. This resulted in four continuous relative abundance variables (one for each bacterial group) per sample, which sum to 1.0 for each sample. We subsequently converted these bacterial group relative abundances into estimated concentrations (again four continuous variables, one for each bacterial group) making use of the BactQuant results as described above. The BactQuant data were also used to determine estimated concentrations of taxa of interest and the total bacterial load in each sample.

*Example to illustrate VMB types and bacterial groups*

A sample containing a 30% relative abundance of *L. iners*, 30% other lactobacilli, and 40% BV-anaerobes, would have been assigned to the LA VMB type. If this sample contained one million 16S rRNA genes per μl, and each of the species included in the sample only contained one 16S rRNA gene copy, it would contain estimated concentrations of [30%+30%] x one million = 600,000/μl lactobacilli, [40%] x one million = 400,000/μl BV-anaerobes, 0/μl pathobionts, and 0/μl other bacteria.

#### Statistical analyses

*Software used*

Clinical and laboratory data we entered into a Microsoft Excel spreadsheet. Statistical analyses, bar charts, and scatter plots were done in STATA version 15.1 (StataCorp, College Station, TX, USA). The heatmap in Figure A1 was made using the *gplots* package in R version 3.5.1 (R core team, 2015).^20^ The Venn diagrams in Figure A2 were made using a website produced by Meta-Chart.^21^

*Sample size calculation*

No formal sample size calculations were done because this was a secondary exploratory analysis.

*Statistical analyses performed*

These are described in the manuscript. Missing data were not imputed.

*Multiple comparisons*

We performed a large number of comparisons and this might generate some false-positive results. However, the VMB composition variables overlapped considerably (for example CSTs, VMB types, and *Lactobacillus* groups). Therefore, findings using one classification system would be expected to correlate with findings using other but similar classification systems. We focused on such consistencies, rather than individual statistically significant findings, and therefore decided not to adjust for multiple comparisons.

#### Crown initiative

The CROWN initiative has defined a core outcome set for PTB research.^21^ We reported on PPROM, birthweight, and gestational age at birth. There were no cases of maternal mortality. The remainder of the core outcome set is not relevant to this study. During the course of the research, the PREBIC consortium^22^ also made recommendations for a minimum dataset for research on the VMB in PTB.^23^ Despite the recommendations not having been formulated during data collection, we present 24/26 of the essential criteria and 11/18 of the desirable criteria (Table A2).

#### Patient and public engagement

Patients who had previously received care at Liverpool Women’s Hospital preterm birth prevention clinic had contributed to the successful funding bid to establish the Harris-Wellbeing Preterm Birth Research Centre at Liverpool Women’s Hospital. A part of this funding was used for the “Development of novel biomarkers for prediction of preterm labour in a high-risk population” study, of which the current project is a sub-study. After the successful funding bid, in 2015, a formal Harris-Wellbeing Preterm Birth Research Centre Patient and Public Involvement (PPI) group was formed. The group consisted of highly motivated parents, the majority of whom had previously experienced a PTB. The group met approximately 3 times/year over the course of participant recruitment and helped guide the research team on the practicalities of recruitment. This included helping develop the participant information leaflet, optimal language to use with regards to the process of obtaining high vaginal swabs and optimal ways to approach the low-risk participants. The group has not been involved in data analysis but the study results were explained to them. The hard-work and dedication of the group was recognised when they were finalists in the University of Liverpool PPI group awards 2018.

### Supplementary Figures

**Figure A1: Heatmap of the bacterial groups and 13 most abundant amplicon sequence variants**

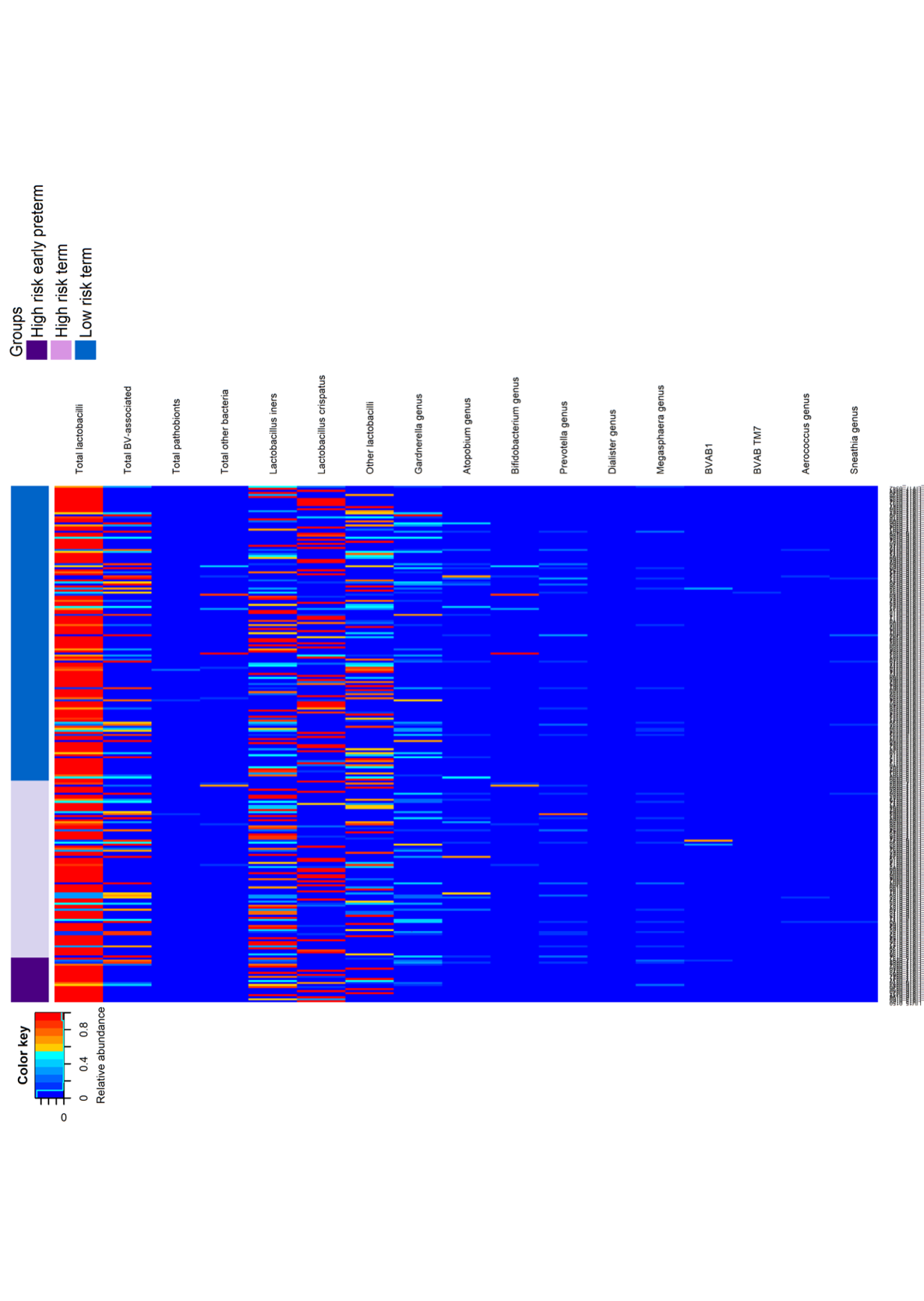

Abbreviations: BV, bacterial vaginosis; BVAB1, BV-associated bacterium type 1; BVAB TM7, BV-associated bacterium (phylum TM7).

This heatmap depicts all samples (*n* = 254) on the x-axis: one column represents one sample. The top bar depicts pregnancy outcome group. The next four bars show relative abundances of the four mutually exclusive bacterial groups, and the bottom 13 bars relative abundances of the 13 most abundant amplicon sequence variants.

**Figure A2: Associations between VMB characteristics and recurrent early sPTB/PPROM**

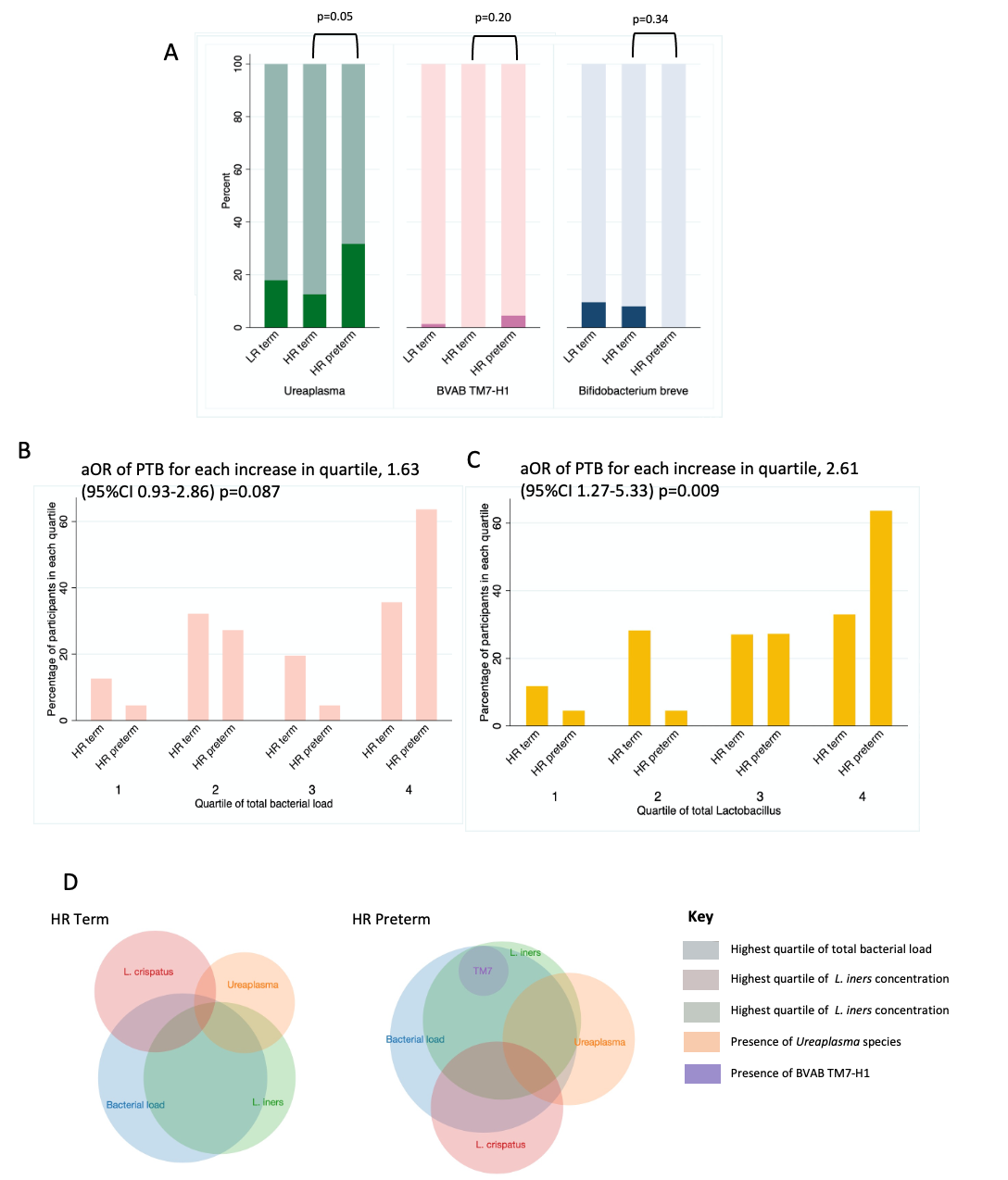

Abbreviations: LR term, low-risk pregnancies with birth ≥39^+0^ weeks; HR term, high-risk pregnancies with birth ≥ 37^+0^ weeks; HR preterm, high-risk pregnancies with spontaneous preterm birth (SPTB) or premature prelabour rupture of membranes (PPROM) <34^+0^ weeks.

**A)** Bacterial taxa of interest presence in percentage of LR term, HR term, and HR preterm women. Dark colour= taxa present; light colour= taxa not present. **B-C)** Percentage of women in each quartile of total bacterial load (B) and total estimated *Lactobacillus* concentration (C) for the HR term and HR preterm groups. Quartiles were generated using LR term cohort distributions. Odds ratios are adjusted for BMI, cervical surgery, and smoking. **D)** Venn diagrams showing the co-location of VMB characteristics that were positively associated with early preterm birth recurrence within the high-risk cohort. 17/22 (77%) of HR preterm and 44/87 (50.6%) of HR term participants had at least one VMB characteristic associated with early sPTB/PPROM recurrence. The highest quartile of bacterial load had complete overlap with highest quartile of total *Lactobacillus* concentration, and so only total bacterial load is shown.

Supplementary TableS

**Table A1: Previous studies using vaginal microbiome sequencing to assess the associations between VMB composition and PTB**

| **Part 1: Study Characteristics** | | Romero  2014^22^ | DiGiulio  2015^23^ | Subramania  2016^24^ | Nelson  2016^25^ | Kindinger  2017^26^ | Callahan  2017^27^ | Stout  2017^28^ | Freitas  2018^18^ | Tabatanaei 2019^29^ | Brown  2019^30^ | Elovitz  2019^19^ | Fettweis  2019^7^ |
| --- | --- | --- | --- | --- | --- | --- | --- | --- | --- | --- | --- | --- | --- |
| Population | Number of term birth participants | 72 | 34 | 20 | 27 | 184 | 85 | 53 | 170 | 356 | 36 | 432 | 90 |
|  | Number of PTB participants | 18 | 15 | 20 | 13 | 44 | 50 | 24 | 46 | 94 | 60 | 107 | 45 |
|  | Inclusion of sPTB cases | **✓** | **✓** | **✓** | **✓** | **✓** | **✓** | **✓** | **✓** | **✓** | ✘ | **✓** | **✓** |
|  | Inclusion of PPROM cases | **✓** | **✓** |  | **✓** | ✘ | **✓** | **✓** | **✓** |  | **✓** | **✓** | **✓** |
|  | Inclusion of medically indicated PTB | ✘ | **✓** | ✘ | ✘ | ✘ | **✓** | **✓** | ✘ | ✘ | ✘ | ✘ | ✘ |
|  | Percent of PTB under 34 weeks | 100% | 20% | 100%  (<35) |  | 52% | 58% | 33% | 13%  (<32) | 18% |  | 75% (<35) |  |
|  | Gestation of sampling (weeks) | 6-birth | 10-birth | 21-25^+6^ | 9-24 | 16-34 | 6-birth | 6-36 | 11-16 | 8-13^+6^ | 6-36^+6^ | 16-28 | 6-birth |
| Laboratory methods | 16s rRNA sequencing | **✓** | **✓** | **✓** | **✓** | **✓** | **✓** | **✓** | ✘ | **✓** | **✓** | **✓** | **✓** |
|  | Variable region sequenced | 1-3 | 3-5, 4 | 4 | 4 | 1-3 | 4 | 1-3,3-5 |  | 4 | 1-2 | 3-4 | 1-3 |
|  | qPCR of 16s rRNA gene | ✘ | ✘ | ✘ | ✘ | ✘ | ✘ | ✘ | **✓** | ✘ | ✘ | **✓** | ✘ |
|  | Alternate NGS of eukaryotic DNA* | ✘ | ✘ | ✘ | ✘ | ✘ | ✘ | ✘ | **✓** | ✘ | ✘ | ✘ | **✓** |
| Ethnicity | Caucasian/White/European ancestry | 6% | 59% | 50% | 0% | 65% | 23% |  | 60% | 73% | 51% | 21% | 14% |
|  | Black/African American | 87% | 4% | 50% | 100% | 19% | 59% | 69% | 2% | 7% | 23% | 75% | 78% |
|  | Asian |  | 12% | 0% | 0% | 17% | 4% |  | 14% | 4% | 26% |  |  |
|  | Other/mixed | 6% | 25% | 0% | 0% | 0% | 13% | 31% | 24% | 16% | 0% | 4% | 7% |
|  | Control for confounders (either statistically or by matching) | ✘ | ✘ | ✘ | Omitted previous  antibiotics | Age, BMI, ethnicity | ✘ | GA at sampling | ✘ | GA at sampling | Age, BMI, ethnicity | Ethnicity | Age, race, income |
| Relationship between VMB and PTB suggested | | ✘ | **✓** | ✘ | **✓** | **✓** | **✓** | **✓** | **✓** | **✓** | **✓** | **✓** | **✓** |

Abbreviations: BMI, body mass index; GA, gestational age; NGS, next generation sequencing; PPROM, preterm prelabour rupture of membranes; qPCR, quantitative PCR; sPTB, spontaneous preterm birth.

*Alternate NGS of eukaryotic DNA was performed by cpn60 universal target sequencing^35^ and shotgun metagenomic/metatranscriptomic sequencing^9^. **✓**=’yes’, ✘=’no’. Grey=not available/not applicable.

| **Part 2: Findings associated with PTB** | | Romero  2014^22^ | DiGiulio  2015^23^ | Subramania  2016^24^ | Nelson  2016^25^ | Kindinger  2017^26^ | Callahan  2017^27^ | Stout  2017^28^ | Freitas  2018^18^ | Tabatanaei 2019^29^ | Brown  2019^30^ | Elovitz  2019^19^ | Fettweis  2019^7^ |
| --- | --- | --- | --- | --- | --- | --- | --- | --- | --- | --- | --- | --- | --- |
| Global sample assessments | Species richness |  |  |  | ⇔ | ⇔ |  | **↑** | **↑** |  | **↑** |  |  |
|  | Species diversity (alpha diversity) | ⇔ | **↑** | ⇔ | ⇔ | ⇔ |  | **↑** | **↑** | ⇔ | ⇔ | ⇔ | **↑** |
|  | Community state types | ⇔ | CST IV **↑** |  | ⇔ | CST III ↑, I ↓ |  |  | ⇔ | CST IV **↑** |  | CST IV **↑** | CST I ↓ |
|  | *Lactobacillus* relative abundance group |  |  |  |  | ⇔ | Low *lactos* |  |  |  | Low *lactos* |  |  |
|  | Instability |  |  |  |  |  | **↑** | **↑** |  |  | **↑** |  | ⇔ |
|  | Total bacterial load |  |  |  |  |  |  |  | **↑** |  |  | ↓ |  |
| Relative abundance of specific taxa of interest | *Lactobacillus* species | ⇔ | ↓ |  | ⇔ | ⇔ | ↓ | ⇔ | **↑** |  | ↓ |  |  |
|  | *Lactobacillus crispatus* | ⇔ |  |  |  | ↓ | ↓ | ⇔ | ⇔ | ↓ | ⇔ | ⇔ | ↓ |
|  | *Lactobacillus iners* | ⇔ |  |  |  | **↑** | ⇔ | ⇔ | ⇔ | ↓ | ⇔ | **↑** | ⇔ |
|  | *Lactobacillus jensenii* | ⇔ |  |  |  | ⇔ | ↓ | ⇔ | ⇔ | ⇔ | ⇔ | ⇔ | ⇔ |
|  | *Lactobacillus gasseri* | ⇔ |  |  |  | ⇔ | ↓ | ⇔ | ⇔ | ↓ | ⇔ | ⇔ | ⇔ |
|  | *Aerococcus* | ⇔ |  |  | ↓ | ⇔ | **↑** | ⇔ | ⇔ | ⇔ |  | ⇔ | **↑** |
|  | *Atopobium vaginae* | ⇔ |  |  |  | ⇔ | **↑** | ⇔ | ⇔ | ⇔ | ⇔ | **↑** | ⇔ |
|  | *Bifidobacterium breve* |  |  |  | ⇔ | ⇔ | ⇔ | ⇔ | ⇔ | ↓ | ⇔ | ⇔ |  |
|  | *Clostridiales* BVAB2 | ⇔ |  |  |  |  | ⇔ | ⇔ |  |  |  | ⇔ | **↑** |
|  | *Dialister* | ⇔ |  |  | ⇔ | ⇔ | **↑** | ⇔ | **↑** | ⇔ | **↑** | ⇔ | **↑** |
|  | *Gardnerella vaginalis* | ⇔ | **↑** |  | ⇔ | ⇔ | **↑** | ⇔ | ⇔ | ⇔ | ⇔ | ⇔ | ⇔ |
|  | *Lachnospiracea* BVAB1 | ⇔ |  |  |  |  | ⇔ | ⇔ |  |  |  | ⇔ | **↑** |
|  | *Mageeibacillus indolicus* |  |  |  |  |  |  |  |  |  |  | **↑** |  |
|  | *Megasphaera* | ⇔ |  |  | ⇔ |  | ⇔ | ⇔ | **↑** | ⇔ | ⇔ | **↑** | **↑** |
|  | *Mobiluncus* |  |  |  |  | ⇔ | **↑** | ⇔ | **↑** |  |  | **↑** |  |
|  | *Mycoplasma* |  |  |  |  | ⇔ | **↑** | ⇔ | **↑** |  |  | ⇔ | ⇔ |
|  | *Parvimonas* | ⇔ |  |  | ⇔ | ⇔ |  | ⇔ | **↑** |  | **↑** |  | **↑** |
|  | *Peptoniphilus* |  |  |  |  | ⇔ | **↑** | ⇔ | **↑** | ⇔ | **↑** | ⇔ |  |
|  | *Prevotella* | ⇔ |  |  | ↓ | ⇔ | **↑** | ⇔ | **↑** | ⇔ | **↑** | ⇔ | **↑** |
|  | *Porphyromonas* species |  |  |  | ⇔ |  |  | ⇔ | **↑** |  |  | **↑** |  |
|  | *Sneathia* species | ⇔ |  |  | ↓ | ⇔ | ⇔ | ⇔ |  | ⇔ | ⇔ | **↑** | **↑** |
|  | *Streptococcus* species | ⇔ |  |  | ⇔ | ⇔ | **↑** | ⇔ | **↑** | ⇔ | **↑** | ⇔ | ⇔ |
|  | BVAB TM7-H1 | ⇔ |  |  |  |  | ⇔ | ⇔ |  |  |  |  | **↑** |
|  | *Ureaplasma* species | ⇔ | **↑** |  |  | ⇔ | ⇔ | ⇔ | **↑** |  | ⇔ | ⇔ | ⇔ |

Abbreviations: BVAB1, BV-associated bacterium type 1; BVAB TM7, BV-associated bacterium (phylum TM7); Lactos, Lactobacillus species; PTB, preterm birth; VMB, vaginal microbiota.

VMB characteristics associated with PTB in previous studies. ↓=reduced in PTB cases compared to term; ↑=increased in PTB cases compared to term; dark grey= not assessed or not available; ⇔ on white background = tested and no difference clearly shown in manuscript and/or supplementary materials; ⇔ on light grey background= testing is inferred within manuscript and/or supplementary material and no difference shown but statistical tests not shown and/or level of assessment is unclear.

**Table A2: Additional participant characteristics by pregnancy risk group and outcome**

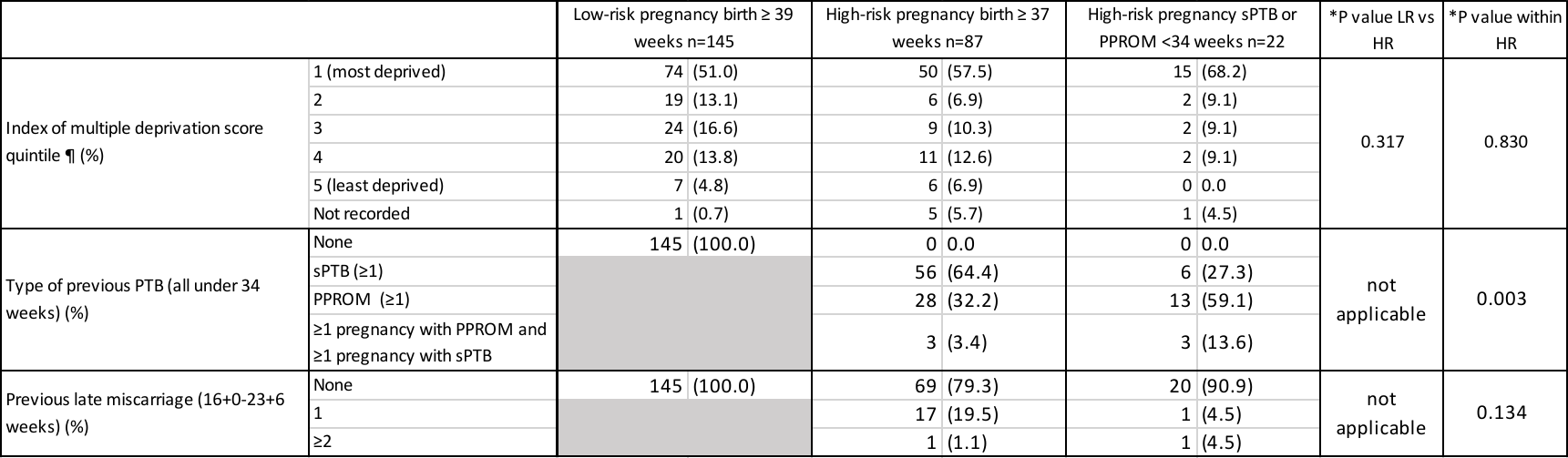

Abbreviations: LR, low-risk pregnancy; HR, high-risk pregnancy; PPROM, preterm prelabour rupture of membranes; sPTB, spontaneous preterm birth.

*P-values by Fisher’s exact test.

¶The index of multiple deprivation was obtained using the woman’s home postcode on the UK government website.^31^ The index ranks every neighbourhood in England from 1 (most deprived) to 32844 (least deprived).^32^ It is a collective score summarising income deprivation, employment deprivation, health deprivation and disability, education skills and training deprivation, barriers to housing and services, living environment deprivation, and crime.

**Table A3: Relative abundances of bacterial groups and taxa of interest by pregnancy risk cohort and outcome**

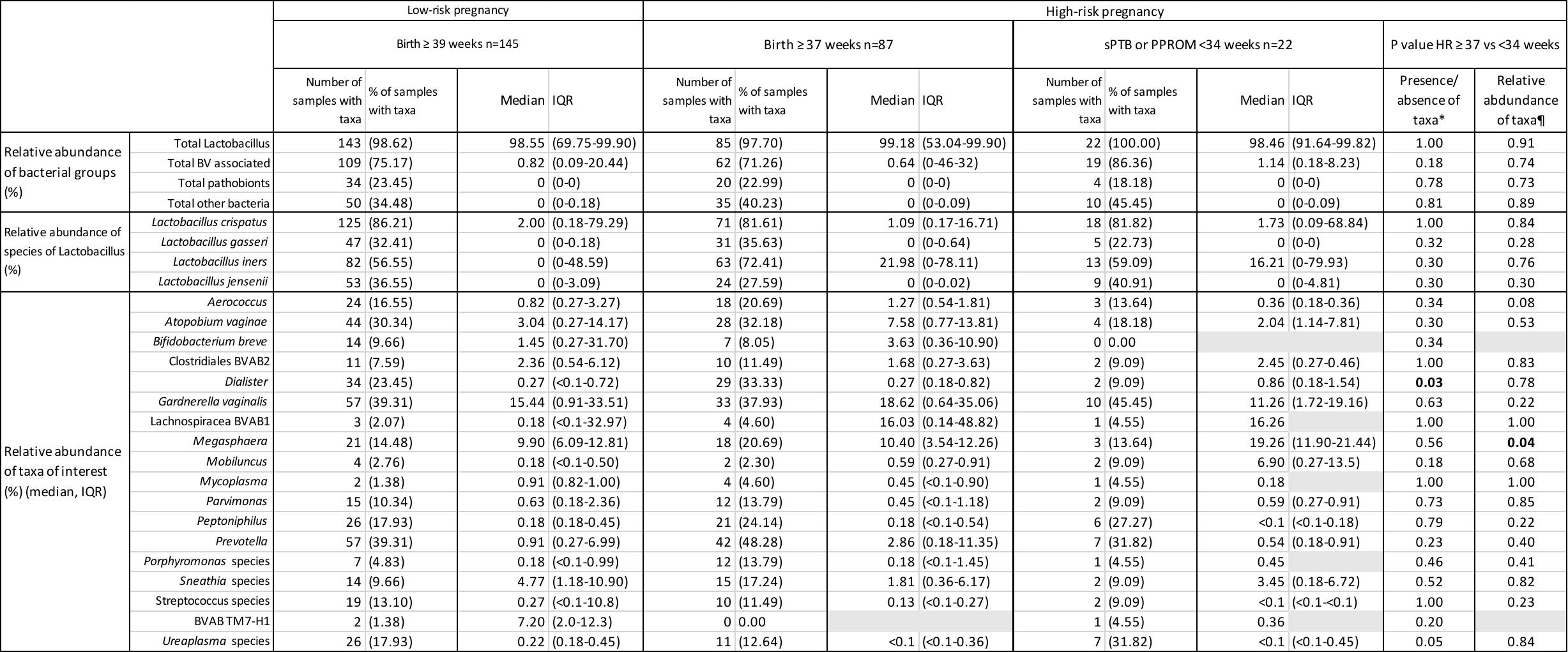

Abbreviations: LR, low-risk pregnancy; HR, high-risk pregnancy; PPROM, preterm prelabour rupture of membranes; sPTB, spontaneous PTB. Grey box, value not applicable due to no sample fitting this criterion.

*P values by Fisher’s exact test for presence/absence of bacterial group or taxa.

¶P values by Mann-Whitney U test for relative abundance. Test of significance based on relative abundance includes all participants for relative abundance of bacterial groups and species of lactobacillus but is limited to only those participants with each taxon present for the taxa of interest (to avoid skewing of data in cases of rare taxa).

**Table A4: P-values for mean bacterial load comparisons between VMB types in high-risk women**

The p-values (by Mann-Whitney U test) in the table below are for mean bacterial load comparisons between VMB types in high-risk women. The mean bacterial loads for each VMB type are reported in Table 3 in the manuscript.

|  | **N** | **Lcr** | **Li** | **Lo+BL** | **LA+BV** |
| --- | --- | --- | --- | --- | --- |
| ***L. crispatus*-dominated (Lcr)** | 23 |  | 0.475 | 0.946 | 0.008 |
| ***L. iners*-dominated (Li)** | 36 | 0.475 |  | 0.361 | 0.033 |
| **Other lactobacilli- or *Bifidobacterium*-dominated (Lo+BL)** | 22 | 0.946 | 0.361 |  | 0.007 |
| **Lactobacilli with anaerobes or BV (LA+BV)** | 28 | 0.008 | 0.033 | 0.007 |  |

VMB type definitions: *L. iners*-dominated (Li; ≥75% lactobacilli with *L. iners* the most common); *L. crispatus* (Lcr; ≥75% lactobacilli with *L. crispatus* the most common); other lactobacilli-dominated or *Bifidobacterium-*dominated (Lo+BL; either≥75% lactobacilli with *L. jensenii* or *L. gasseri* the most common, or ≥50% *Bifidobacterium*); and either lactobacilli and anaerobes (LA; 25%- 75% lactobacilli and the remainder anaerobes) or a mixture of BV-anaerobes (BV; ≥75% BV-anaerobes).

**Table A5: PREBIC consortium essential and desirable criteria of minimum PTB dataset**^8^

| **Item required** | **Location within manuscript, or reason for absence** |
| --- | --- |
| **Essential** |  |
| Age | Table 1 |
| Race/ethnicity | Table 1 |
| Parity | Table 1 |
| BMI | Table 1 |
| Smoking status | Table 1 |
| History of sexual transmitted infection | Information not collected |
| History of PTB | Table 1 |
| Indication of previous PTB | All previous PTB either sPTB <34 weeks or after PPROM <34 weeks. Details in Table A2 |
| Information on included singleton/multiple pregnancy | All singleton pregnancies (main document, methods) |
| Exclusion of other complications in pregnancy leading to PTB | Study team excluded other causes of PTB (Figure 1 and main document, methods) |
| Information on use of antibiotics before sampling | Table 1 |
| Information on use of antibiotics after sampling | Data not collected |
| Adequate assessment of gestational age | All pregnancies had ‘dating’ scan prior to 14 weeks gestation (Appendix A, supplemental methods) |
| Gestational age at sampling | Table 1 |
| Single or longitudinal sampling | Main document, methods |
| Information on primary swab | HydraFlock standard tapered swabs, (Medical Wire and Equipment, Corsham, England) |
| Sample location | All swabs taken and stored at Liverpool Women’s Hospital, UK |
| Information on primers used | Appendix A ‘PCR amplification and 16S rRNA gene sequencing’ section |
| Range of bacteria covered by primers | Good coverage of vaginal microbiota, as described by Van der Pol *et al*^33^ |
| Attrition record | Figure 1 |
| Statement of outcome measures | Main document, Methods and Appendix A ‘Vaginal microbiota characterisation’ |
| Definition of PTB | Main document ‘Study population’ |
| Indication for PTB of index pregnancy | Table 1 |
| Stratification of PTB by phenotype | Attempted, but as described in limitations no difference identified and data presented as a whole to improve readability and statistical power |
| Races/ethncities analysed separately | Not applicable because over 90% of population white. |
| Analysis of Lactobacilli to species level | Tables 2,3 A3 |
| Other interventions for PTB excluded (eg cerclage, pessary) | Women with interventions for PTB not eligible for recruitment. Participants with interventions after VMB analysis are included, as discussed in limitations section |

| **Desirable** |  |
| --- | --- |
| Marital status | Not collected |
| Socioeconomic status | Table A2 |
| Alcohol use | Not collected |
| Substrance use | Not collected |
| Gestational age of previous PTB | All previous PTB were sPTB or PPROM 16^+0^ -33^+6^ weeks |
| History of late miscarriage | Table A2 |
| History of LLETZ | Table 1 |
| Date of last sexual intercourse prior to sampling | Appendix A “Participant recruitment” |
| Reported history of douching | Not collected |
| Self collected or physician collected samples | Physician collected, main document methods |
| Measurement of cervical length | Table 1 |
| Use of fetal fibronectin | Table 1 |
| Measurement of pH | Not collected |
| Simultaneous cultivation | Not performed by research team |
| Objective measurement of BV by microscopy | Not performed by research team |
| Consent to use specimens | Consent documented for ‘gifting’ of samples to other ethically approved research |
| Other subtypes of PTB (< 34 weeks, <28 weeks) | Primary outcome sPTB or PPROM 16^+0^-33^+6^ weeks. Insufficient sample size to justify further subtypes |
| Quantitative/qualitative analysis | Performed comprehensively. Tables 2 & 3. |

Note: these recommendations were published after we had already completed study recruitment.
